# How lethal is the novel coronavirus, and how many undetected cases there are? The importance of being tested

**DOI:** 10.1101/2020.03.27.20045062

**Authors:** Ugo Bastolla

## Abstract

There is big concern for estimating the lethality and the extent of undetected infections associated with the novel coronavirus SARS-CoV2 outbreak. While detailed epidemiological models are certainly needed, I suggest here an orthogonal approach based on a minimum number of parameters robustly fitted from the cumulative data easily accessible for all countries at the John Hopkins University database that became the worldwide reference for the pandemics. I show that, after few days from the beginning of the outbreak, the apparent death rate can be extrapolated to infinite time through regularized regression such as rescaled ridge regression. The variation from country to country of these extrapolated death rates appears to depend almost only (*r*^2^ = 0.91) on the ratio between performed tests and detected cases even when the apparent instantaneous lethality rates are as different as 9% in Italy and 0.4% in Germany. Extrapolating to the limit of infinite number of tests, I obtain a death rate of 0.012 ± 0.012, in agreement with other estimates. The inverse relationship between the extrapolated death rate and the intensity tests allows estimating that more than 50% of cases were undetected in most countries, with more than 90% undetected cases in countries severely hit by the epidemics such as Italy. Finally, I propose to adopt the ratio between the cumulative number of recovered and deceased persons as an indicator that can anticipate the halting of the epidemics.

---

The pandemic spread of the novel coronavirus SARS-CoV2 is causing thousands of fatalities, creating a tremendous threat to global health [1]. In this situation, the society is strongly concerned by the lethality and the true extension of the pandemics, see for instance [2,3]. In the media, but also in some non-specialist scientific circles, it is frequent to find that lethality is estimated as the cumulative number of deaths divided by the cumulative number of confirmed cases, data that are easily accessible to everyone in the internet. This quantity changes rapidly and systematically both in time and in space, generating doubts and concerns regarding its interpretation. Of course, epidemiologists can estimate the lethality rate in a less biased way with additional data on the dates in which the infections of the persons that die or recover were detected, and better sampling of the population. However, these data are not easily accessible for all countries. Therefore, here I set up to obtain an estimate of the lethality rate and the detection rate only based on the data reported for all countries in the John Hopkins University database [4]. This extrapolation of the data shows that, when the time course of the disease is controlled for, the lethality estimated for all countries for which reliable data are available depends only on the intensity of the performed tests, i.e. the number of tests divided by the number of positive case. Extrapolating to infinite number of tests, I estimate a lethality rate of 0.012 ± 0.012, which is very noisy but consistent with the estimate of 0.01 that is frequently mentioned in the media. Inverting the relationship between apparent death rate and number of tests, it is possible to estimate that in all countries except South Korea and perhaps Germany, at least at the beginning of the spread, the vast majority of positive cases went undetected, with more than 90 percent undetected cases in some countries such as Italy. These cases that went out of the radar and were not isolated are likely to have contributed strongly to the rapid spread of the virus. Finally, I propose to adopt the ratio between the cumulative number of recovered and the cumulative number of deceased persons, as a potential indicator that can anticipate whether the spread of the epidemics is halting.

The ratio between the number of death and the number of confirmed cases is clearly an underestimate of the lethality, because it assumes that all the presently infected people will eventually recover; but it is also an overestimate, because it assumes that all the extant infections are detected. To mitigate the first problem, it is possible to extrapolate the lethality to the infinite time when all the infected cases are resolved. The asymptotic lethality rate *d* is the fraction of present confirmed cases (*N*) that are already deceased (*D*) plus the presently infected persons *I* = *N* − *R* − *D* that will eventually die, *d* = (*D* + *d*_1_*I*)*/N* ⇒ *D/N* = *d* − *d*_1_(*N* − *R* − *D*)*/N* (here *R* is the cumulative number of recovered persons). If we assume that *d*_1_ is constant, we can estimate the parameters *d* and *d*_1_ through linear regression of the apparent death rate *D/N* versus the infected fraction 1 − (*R* + *D*)*/N*. The assumption of constant *d*_1_ is not realistic in the initial stage of the epidemics but it becomes more and more accurate as time passes, as the data from China shows. A technical point: I regularize the linear regression with the method of rescaled ridge regression [5]. This regularization produces accurate fits while limiting the values of the fitted parameters. Without regularization, the extrapolated death rate *d* would be rather large, while the assumption of constant *d*_1_ is expected to lead to overestimate *d*, in particular in the initial stages of the epidemics. The fitted value of *d*_1_ is similar to, but always smaller than, *d*, which appears reasonable: In fact, the persons that die first belong to the classes with higher death rate, such as the elder and the sick.

Figure 1 shows the curves of *D/N* as a function of the infected fraction. These curves are reasonably linear for large time for most countries. The apparent death rate *D/N* strongly depends on the number of performed tests, as I shall show below, and since this number is not constant, as for instance data from Italy show. Therefore, I fit the lethality rate at the time *t*=12 March at which data on the tests are available for most countries [6] (solid lines in Fig. 1). From these fits, I extrapolate the parameter *d* at the date of 12 March, provided that two conditions hold: at *t* the ratio between cumulative recoveries and cumulative deaths *R/D* is at least 0.9 and *D*(*t*) ≥ 10. When these conditions are not met the asymptotic death rate *d* is clearly overestimated. If the above conditions are not met, I perform a regularized fit over the whole time range (dashed lines in Fig. 1), and again I accept the result only if *R/D >* 0.9 and *D* ≥ 10 at the last available time. For only seven countries I could find data on the number of tests and at the same time I could extrapolate the death rate fulfilling the above conditions: Japan, South Korea, Malaysia, Italy, Spain, France and Switzerland.

**Figure 1:**
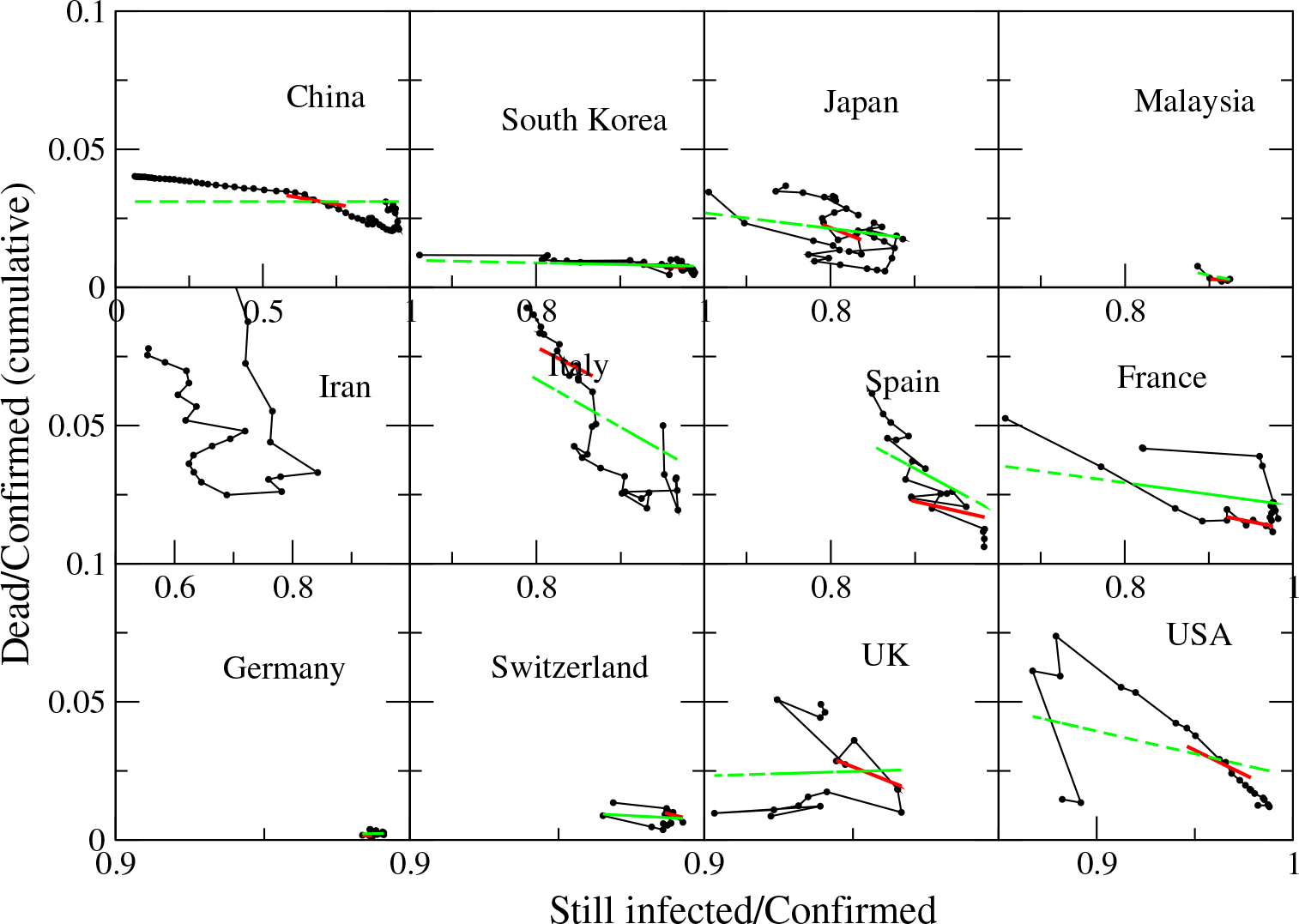
Apparent death rate *D/N* versus the infected fraction 1 −(*R* + *D*)*/N* for several countries with large number of confirmed cases. The thick lines represent the linear fit at *t*=12 March at which data on tests are available. The thin lines represent linear fits that cover all the available range.

The extrapolated *d* is plotted in Fig.2 as a function of the number of confirmed cases *N* divided by the number of tests *T* for the seven countries for which *d* can be reliably estimated. You can see that all countries fall on the same straight line, *d* = *d*_0_ + *a*(*N/T*). This means that the huge apparent differences between the low mortality rates of countries like South Korea or Germany and the high mortality rate of countries like Italy can be fully explained by the time course of the epidemics and the number of tests, without the need of hypothesizing any mutation in the virus. The limit value of the death rate for infinite number of tests can be estimated through linear regression. If the regression is not regularized (dashed line) the estimated death rate is negative. In contrast, with rescaled ridge regression I obtain *d*_0_ = 0.012 ± 0.012, which extrapolates the lethality rate of the coronavirus disease to the situation in which infinite tests are available and all cases have been detected. The fit parameter *a* is *a* = 0.963 and *r*^2^ = 0.906.

**Figure 2:**
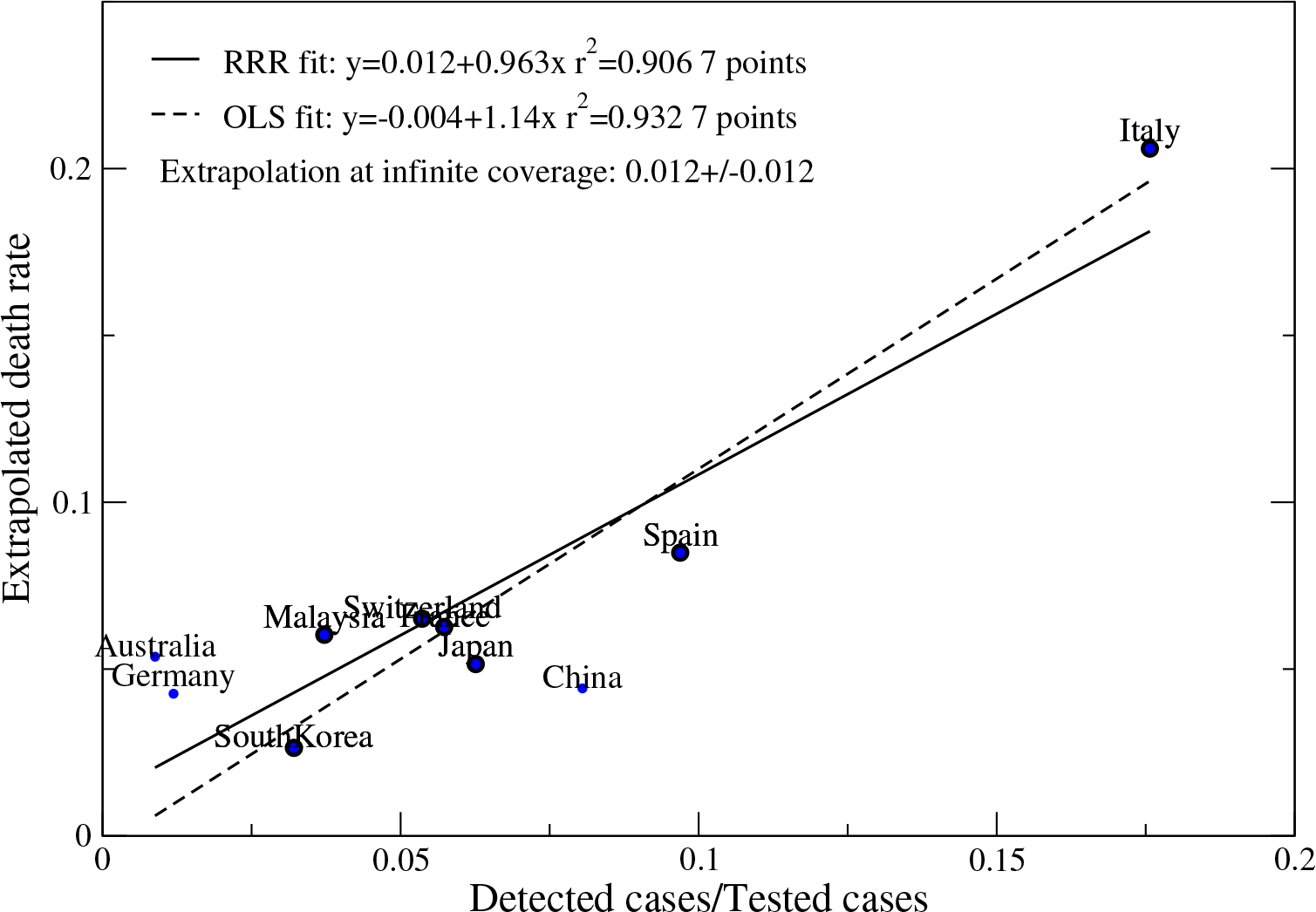
Extrapolated death rate *d* as a function of the number of confirmed cases *N* divided by the number of tests *T* for countries for which *d* can be reliably estimated. The fits extrapolate the death rate for infinite tests. Small dots represent countries whose extrapolated death rates and/or number or tests are not reliable and are not used in the regression, as discussed in the text.

For reference, Fig.2 also reports data for China, Germany and Australia. In the case of China, I could only find the number of tests performed in the Guangdong province and estimated that the number of tests performed in the whole country was 3 times larger. If this estimate is approximately correct, the data suggests that the mortality rate of China is smaller than for other countries, which could be attributed to two factors: (1) The Chinese population is significantly younger (for instance, only 12% of the Chinese population has more than 65 years, compared to 23% for Italy). (2) China is closer to the extinction of the epidemics, where the extrapolation at infinite time becomes quite accurate, while at smaller time the extrapolation probably (and hopefully) overestimates the mortality rate. For Germany, I could only find the number of tests performed in private laboratories and estimated the total number of tests as the double of this number.

Moreover, the fit over the whole time range yielded negative extrapolated death rate, and at 12 of March it was *D*(*t*) = 2, so that the estimate reported in the plot is probably an overestimate. Finally, for Australia it was *D* = 3 at March 12 and *D* = 7 at the last available day. Despite these three data are not reliable, they do not deviate too much from the general trend.

From the above relationship between intensity of tests and apparent death rate we can estimate the fraction of undetected cases, *U* = 1 − *N/M*, where *M* is the true number of infected persons: *d*_0_ = *D/M* = *D/N* − *a*(*N/T*) ⇒ *U* ≡ 1 − *N/M* = 1*/*(1 + (*d*_0_*/a*)(*N/T*)). Using the parameters *d*_0_ and *a* determined from Fig.2, I estimate the fraction of undetected cases for various countries and show it in Fig.3. One can see that this fraction is substantially larger than 50% for most countries, and it is larger than 90% for some countries like Italy.

**Figure 3:**
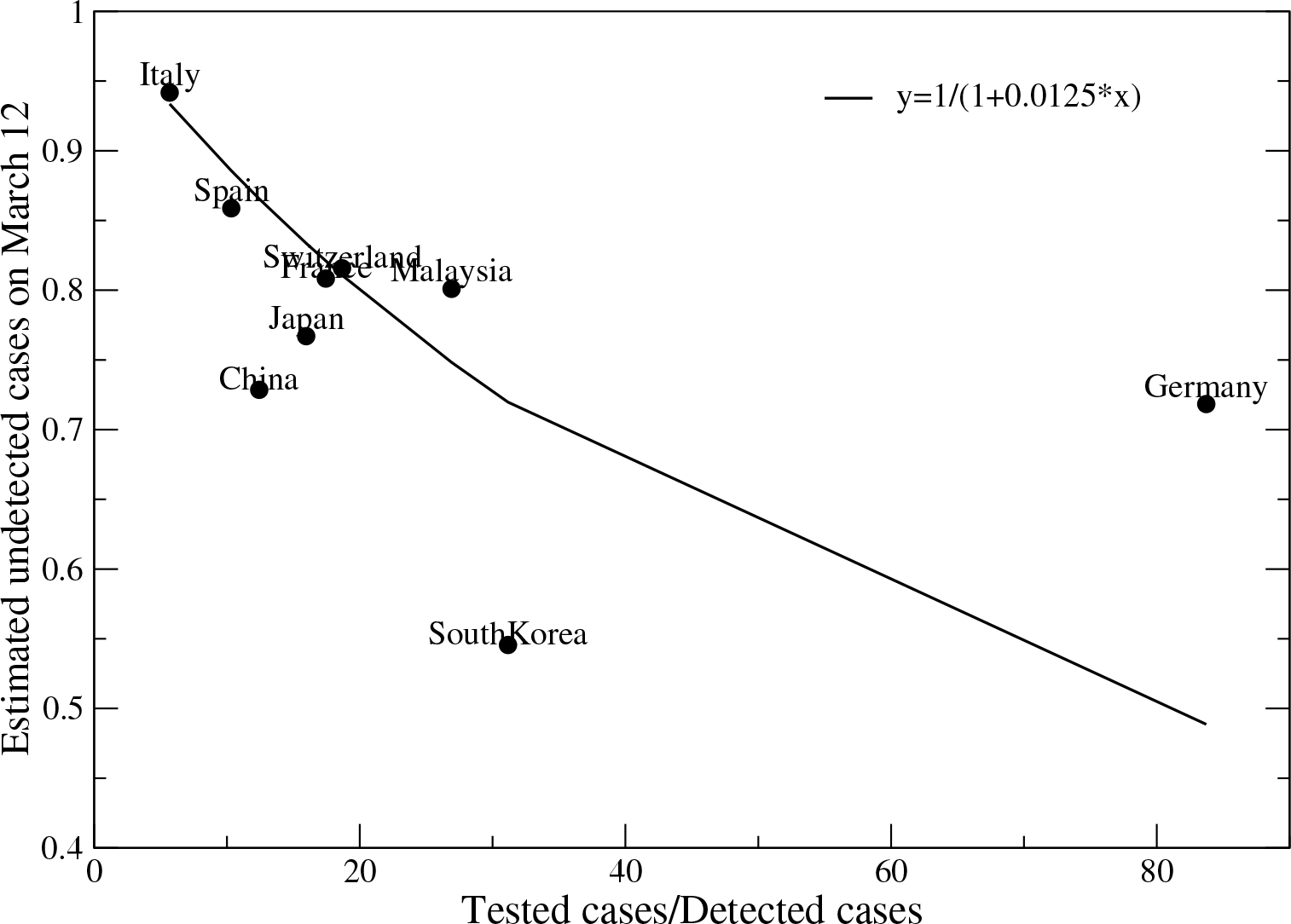
Fraction of undetected cases,*U* = 1 − *d*_0_*/d* versus coverage of confirmed cases *T/N*. The straight line is the relationship expected from the linear fit of the previous figure.

We examine more in detail the effect of the number of tests for the Italian data in Fig.4, using the data made available by its Health Ministery [7]. One can see that the number of tests per confirmed cases decreased with time (A), as the consequence both of the saturation of resources and of the decision to test only syntomatic subjects (March 25th), which was probably unfortunate since it left the majority of cases uncovered. As the number of tests decreased, the apparent instantaneous mortality rate *D/N* increased (B), again in agreement with the linear relationship *D/N* = *δ* + *α*(*N/T*). The non-regularized linear fit produces the unphysical parameter *δ* = −0.0136, whereas rescaled ridge regression yields *δ* = 0.0029 (Fig.4B). Note that this low instantaneous mortality rate only takes into account those that are already dead at time *t* and does not consider the extrapolated mortality. Once again, extrapolated to infinite number of tests, data from Italy are consistent with the much lower apparent mortality rates observed in Germany. As the intensity of tests decreased, the fraction of confirmed cases that were weak enough to be treated at home at first decreased and then increased again (Fig.4C), possibly as the consequence of the almost saturation of the health system that encouraged patients in not too serious conditions to stay at home. Finally, the estimated fraction of undetected cases *U* increased with decreasing *T/N* (Fig.4D). This is of course not surprising, since *U* is computed from *T/N* : The aim of the figure is only to quantify the expected impact of reducing the intensity of tests, which appears to be modest.

**Figure 4:**
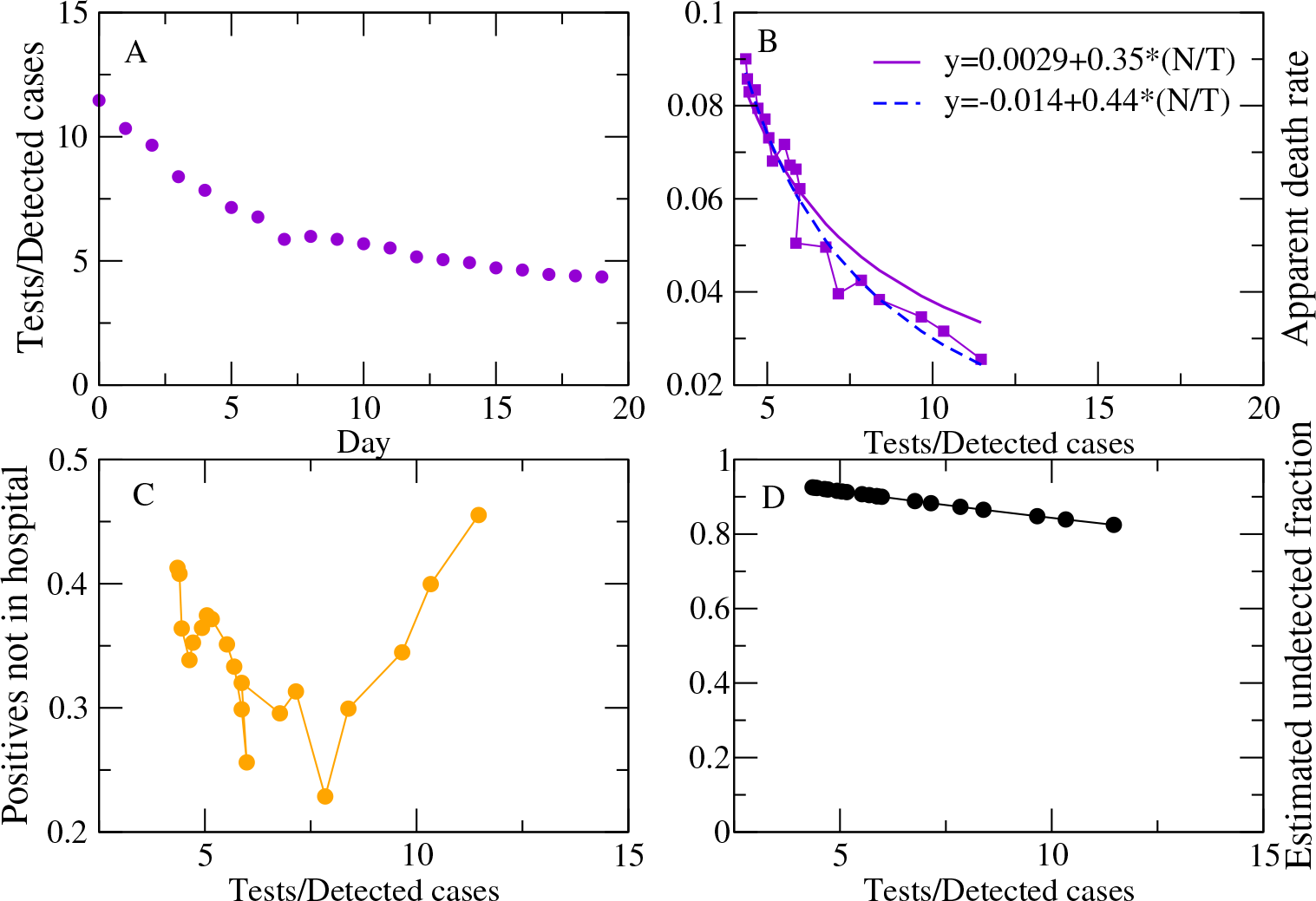
A: The number of tests per confirmed cases decreased with time in Italy. B: The apparent instantaneous mortality rate *D/N* increased when the test intensity *T/N* decreased as *D/N* = *δ* + *α*(*N/T*). We report both the non-regularized fit (dashed) and rescaled ridge regression (solid). C: Fraction of confirmed cases that were weak enough to be treated at home versus *T/N*. D: Estimated fraction of undetected cases versus *T/N*.

Finally, making one step back, Fig.5 shows the ratio between the cumulative number of recovered persons and the cumulative number of deaths, *R*(*t*)*/D*(*t*). This ratio shows complex dynamics that contradicts the simplifying assumption of constant *d*_1_. The data suggest that *R/D* decreases with time when the epidemics spreads exponentially, ultimately reaching a plateau, and it increases with time in the final stages in which the epidemics spreads slowly, which unfortunately is only evident in the data of China, South Korea and perhaps Japan. Thus, the increase of *R/D* might provide an indicator of the halting of the epidemics that is less delayed than the number of deaths alone and less affected by the number of tests than the confirmed cases. Unfortunately, the John Hopkins University database recently stopped the update of the number of recovered persons, possibly because for many countries this data is not sufficiently reliable.

**Figure 5:**
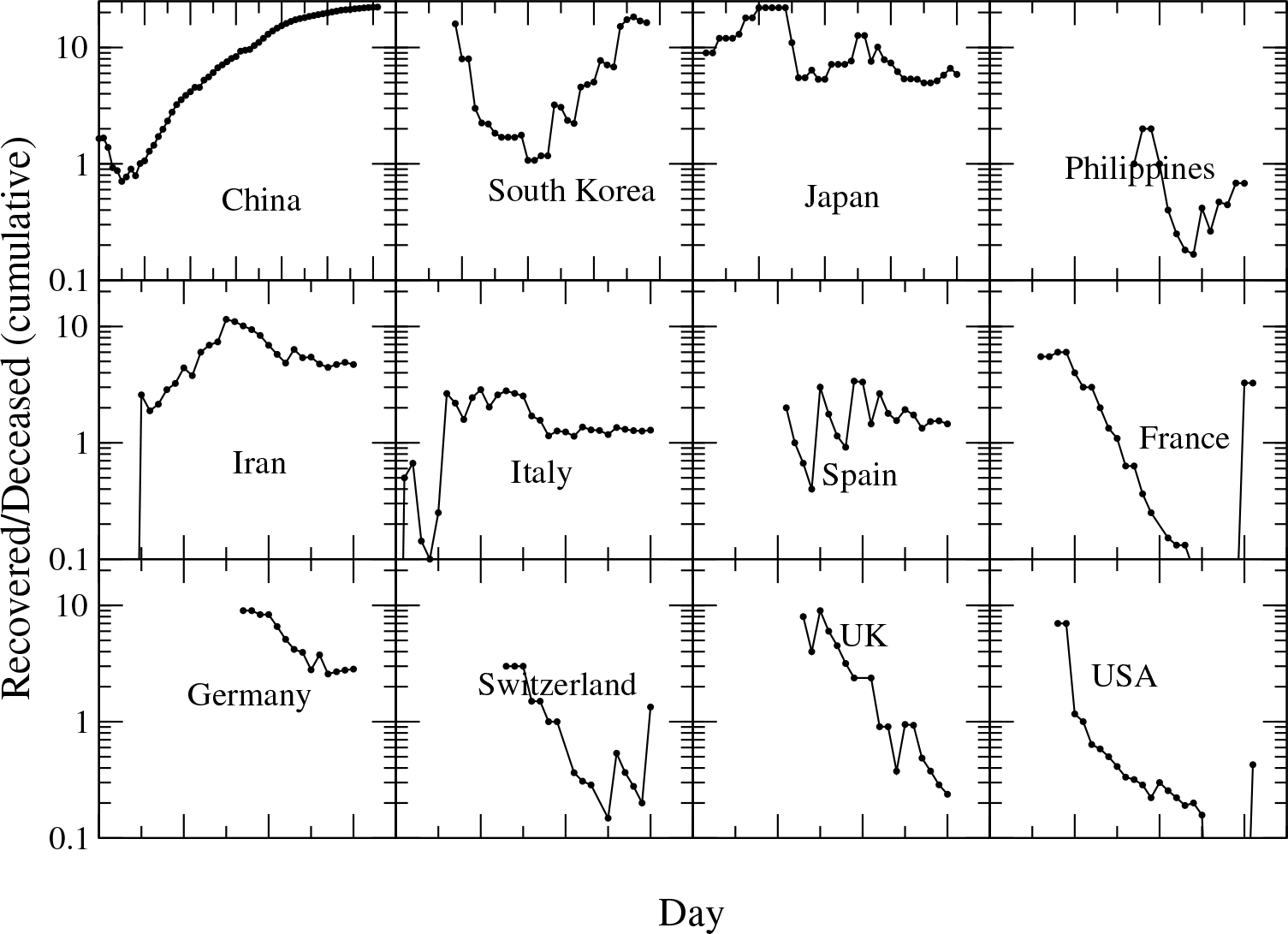
Ratio between the cumulative number of recovered persons and the cumulative number of deaths, *R*(*t*)*/D*(*t*), as a function of time for various countries.

The following is hand-waving reasoning that rationalizes why *R*(*t*)*/D*(*t*) tends to decrease in the exponential phase until reaching a plateau. Denoting by *n*(*t*) the number of people that get the infection at time *t*, and assuming that death takes exactly *t*_*d*_ days, we can estimate the number of people that die at time *t* as *d*(*t*) = *d*_0_*n*(*t* − *t*_*d*_). Analogously, we can estimate the number of recovered people as *r*(*t*) = (1 − *d*_0_)*fn*(*t* − *t*_*r*_), where *t*_*r*_ is the time that it takes to recover and *f* is the fraction of non-lethal infections that are detected and monitored (we assume that this fraction is 1 for infections that result in death). When *n*(*t*) increases exponentially as exp(*t/τ*), the cumulative variables *D*(*t*) = ∫ *d*(*t*) and *R*(*t*) = ∫*r*(*t*) also increase exponentially. So we obtain *R*(*t*)*/D*(*t*) = *f* (1 − *d*_0_)*/d*_0_ exp((*t*_*d*_ − *t*_*r*_)*/τ*) in the exponential phase (here *d*_0_ = 0.012, so the first factor is estimated to be 82*f* that is probably larger than 1 depending on *f*). Note that this expression does not depend on *t*. However, both *t*_*d*_ and *t*_*r*_ are expected to depend on the age group so that *t*_*d*_ decreases for older people and *t*_*r*_ increases. We hypothesize that, in the initial stage, the deaths tend to affect younger persons, who travel more and have more social contacts, while at a later stage they affect more the elders, while people that recover tend to be of the same age throughout the epidemics. These assumptions imply that *t*_*d*_ − *t*_*r*_ decreases with time, producing the observed exponential decrease of *R*(*t*)*/D*(*t*), until the age distribution of people that die and recover becomes stationary. At this point *R/D* becomes approximately constant in the exponential phase, as observed in Fig.5 in the curves from Italy and Spain. Data to test the above hypotheses should be available.

On the other hand, when the epidemics stops to spread exponentially, we expect that *R*(*t*)*/D*(*t*) increases, since *t*_*r*_ tends to be larger than *t*_*d*_, so at the end of the epidemics there is a reservoir of infected people that slowly recover while people that eventually die reach saturation sooner. Therefore, the systematic increase of *R*(*t*)*/D*(*t*) with time may be an early warning that the spread of the epidemics is not spreading exponentially anymore, as suggested by data from China and South Korea.

The main result of this work is that, due to the reduced number of performed tests, the vast majority of the coronavirus infections went undetected in most countries. These undetected infections had the potential to spread freely in the population, giving raise to the rapid exponential phase of the epidemics before the lockout measures.

This result is consistent with recent reports from China [8, 9], and with the high rates of asymptomatic but positive cases observed in Vo Euganeo (Italy) [10], in the cruise ship Diamond Princess [11] and among children [12](see also [13]).

The only countries that were able to keep the fraction of undetected infections low appear to be South Korea and Germany, which explains their better ability to control the epidemics in the initial stages. Unfortunately, in Germany the infection is now spreading exponentially and the tracing of all infections is not anymore a viable option without lockout measures, while the perceived low mortality rate at the initial stage of the infection may act against the adoption of these necessary measures.

## Data Availability

The perl script used for analyzing the data is available upon request

## Acknowledgements

I would like to thank Kiko Llaneras of the Spanish journal El Pais for his insightful articles on the CoVid epidemics that gave me the idea to extrapolate the death rate for infinite testing capacity. David Abia and Pedro Pemau, of the Centro de Biologia Molecular Severo Ochoa (CSIC-UAM) for help with smart-work. People that contributed data on public databases. Giorgio Parisi, Francesca Colaiori, David Abia, Gonzalo Nido, Maria Alieva, Alberto Pascual and Alex Valbuena for useful comments. Most important, thanks to the medical doctors, nurses, police and other public employees that fight against the virus at the risk of their lives.

## References

[1] https://www.who.int/emergencies/diseases/novel-coronavirus-2019

[2] B. Andrino, D. Grasso, K. Llaneras Asíevoluciona la curva del coronavirus en Espaa y en cada autonomí a (2020) https://elpais.com/sociedad/2020/03/17/actualidad/1584436648230452.html (in Spanish)

[3] S. Franchini, E. Marinari, G. Parisi, F. Ricci Tersenghi (2020) Ma quanti sono gli ammalati di Covid-19 in Italia? Scienza in rete, 20/03/2020 (in Italian)

[4] https://github.com/CSSEGISandData/COVID-19/tree/master/cssecovid19data

[5] Bastolla U, Dehouck Y. (2019) Can conformational changes of proteins be represented in torsion angle space? A study with rescaled ridge regression. J Chem Inf Model 59:4929–4941. doi:10.1021/acs.jcim.9b00627

[6] https://ourworldindata.org/covid-testing

[7] https://github.com/pcm-dpc/COVID-19/tree/master/schede-riepilogative/regioni

[8] Wang, C. et al. Preprint at medRxiv https://doi.org/10.1101/2020.03.03.20030593 (2020).

[9] R. Li, S. Pei, B. Chen, Y. Song, T. Zhang, W. Yang, J. Shaman (2020) Substantial undocumented infection facilitates the rapid dissemination of novel coronavirus (SARS-CoV2). Science 10.1126/science.abb3221 (2020)

[10] https://it.euronews.com/2020/03/23/studio-a-vo-euganeo-crisanti-gli-asintomatici-trasmettono-il-coronavirus-covid19

[11] Mizumoto, K., Kagaya, K., Zarebski, A. and Chowell, G. Euro Surveill. 25, 2000180 (2020).

[12] Dong, Y. et al. Pediatrics https://doi.org/10.1542/peds.2020-0702 (2020).

[13] https://www.nature.com/articles/d41586-020-00822-x

